# Improvement of Presenteeism after Treatment of Sleep Disorders in a Tertiary Sleep Center

**DOI:** 10.1101/2025.10.30.25339101

**Authors:** Sakurako Tanno, Teruyoshi Uetani, Atsushi Kikuchi, Hiroshi Shimizu, Yoko Fujino, Tomoko Kira, Yasunori Oka

## Abstract

**Objective:** Patients with sleep disorders who experience excessive daytime sleepiness (EDS) often exhibit reduced work productivity. However, studies investigating whether treatment for sleep disorders improves productivity remain limited. This study aimed to examine the impact of sleep disorder treatment on work productivity, particularly in relation to health-related work limitations, among patients visiting a tertiary sleep center.

**Methods:** Participants were recruited from among patients who had their first consultation at a university-affiliated tertiary sleep center in Japan between 2017 and 2018. A total of 85 patients underwent baseline evaluation using the Epworth Sleepiness Scale (ESS), Pittsburgh Sleep Quality Index (PSQI), Short Form-8 (SF-8), and Work Limitations Questionnaire (WLQ). Thirty-eight participants completed a follow-up evaluation, of whom 23 had complete WLQ data at both time points.

**Results:** At baseline, participants with EDS (ESS ≥ 11) showed significantly lower health-related quality of life, increased sleep disturbances, and reduced work productivity compared with those without EDS. In the longitudinal analysis, all four WLQ subscales (time management, physical demands, mental-interpersonal demands, and output demands) significantly improved after sleep disorder treatment. Among participants with EDS, significant improvements were observed in time management, physical demands, and output demands.

**Conclusions:** Work productivity limitations, as measured by the WLQ, showed improvement following sleep disorder treatment. These findings suggest a potential association between sleep disorder treatment and workplace productivity outcomes. Future studies should further investigate the long-term occupational impact of sleep disorder management.

## INTRODUCTION

Excessive daytime sleepiness (EDS) is a common symptom that can substantially impair the life of affected individuals. A study of 4,722 white-collar workers aged 20 to 59 years in Japan reported a prevalence of EDS, defined as an Epworth Sleepiness Scale (ESS) score of 11 or higher, with 13.3% in women and 7.2% in men [1]. Along with an increased risk of motor vehicle accidents [2] and decreased quality of life [3,4], decreased work productivity is often associated with EDS. For example, in patients with obstructive sleep apnea (OSA), those with both OSA and EDS tended to show significantly lower work productivity than those without EDS [5]. In addition, in a survey of participants diagnosed with narcolepsy, multiple sclerosis, OSA, and depression, as well as those who work shifts, participants with excessive sleepiness showed decreased work productivity compared to those without excessive sleepiness [6]. Impaired work productivity has also been observed in patients with narcolepsy compared to matched controls [7].

Sleep disorders are known to negatively impact workplace productivity and daily functioning. However, the extent to which effective treatment can improve work productivity remains unclear, and data on sleep disorders in clinical settings are particularly limited. We hypothesized that work productivity, as assessed by the WLQ, would improve following sleep disorder treatment in patients with EDS. This study aimed to clarify how presenteeism, defined as decreased work productivity due to health problems, changed after sleep disorder treatment in patients with EDS visiting a tertiary sleep center in Japan. To assess these changes, we used the Work Limitations Questionnaire (WLQ) as the primary outcome measure, with follow-up evaluations conducted at least three months after treatment initiation. The study included patients diagnosed with various sleep disorders, including but not limited to obstructive sleep apnea, insomnia, and narcolepsy, who received guideline-based treatment at the center.

## Materials and Methods

### Participants

This prospective observational study included patients who had their first visit to a university-affiliated tertiary sleep center in Japan between November 2017 and November 2018 and met the age criteria of 18-64 years. Inclusion criteria were as follows:

(1) All new patients attending their initial consultation at the sleep center during the study period were consecutively recruited. (2) Patients aged 18-64 years at the time of their first visit were eligible for inclusion. Exclusion criteria included: (1) those who declined to participate, (2) patients who completed only one visit and did not undergo follow-up, and (3) individuals who had difficulty completing the written questionnaires (e.g., due to visual impairment or inadequate Japanese language skills). Participants were not limited to a specific workplace, industry, or profession, and the study included patients with various sleep disorders, including but not limited to obstructive sleep apnea, insomnia, and narcolepsy.

Of the 215 patients referred to the sleep center during the study period, 158 met the age criteria. Among them, 60 patients declined to participate, seven patients were excluded due to completing only one visit, and two were excluded due to difficulty responding to questionnaires, resulting in 89 patients being enrolled in the study and undergoing baseline evaluation (Figure 1). As this was an observational study, no randomization was applied.

**Figure 1:**
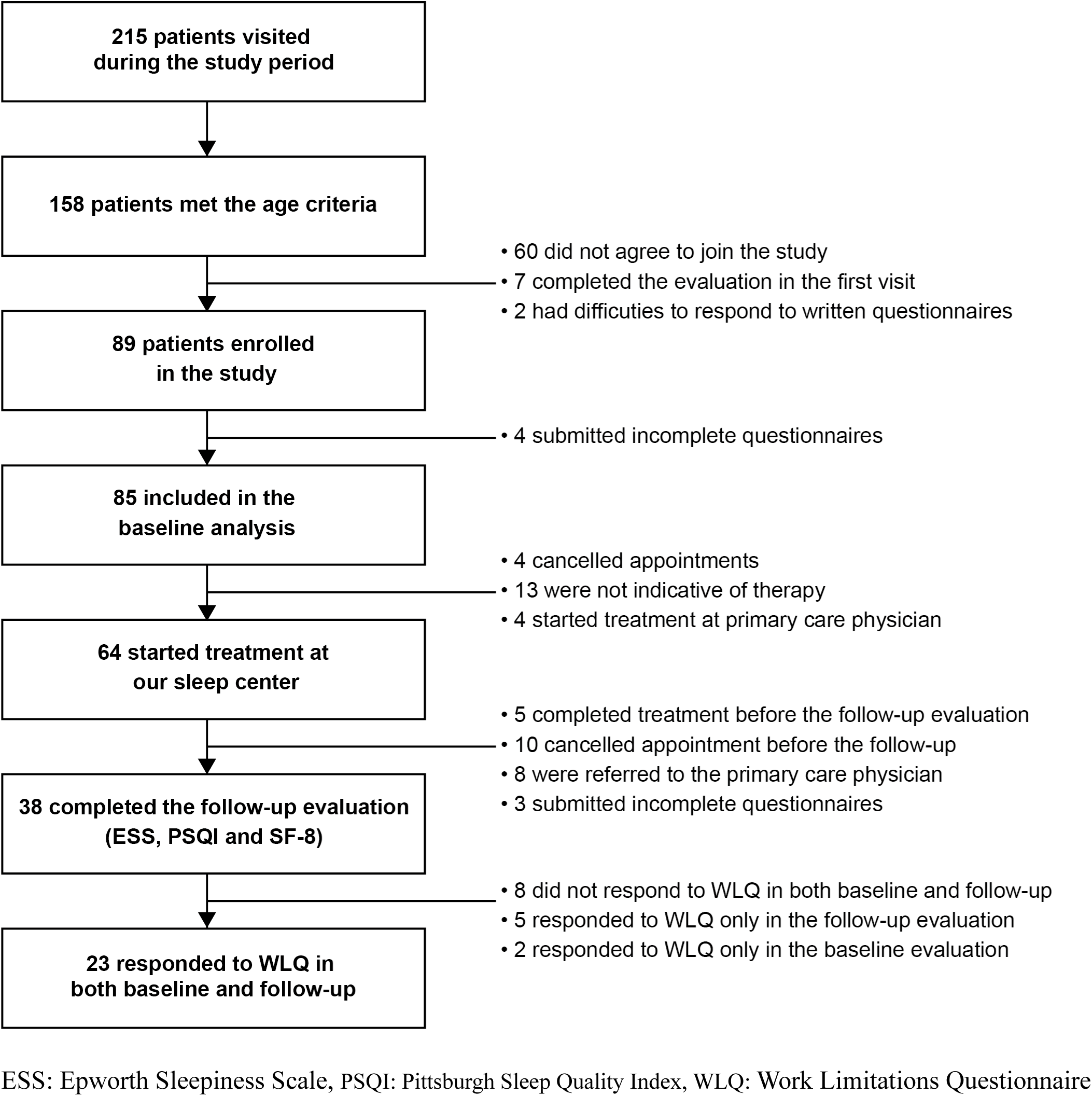
Study protocol of the present study.

All participants provided written informed consent which was approved by the Institutional Review Board of the Ehime University Hospital.

### Data collection and follow-up

At baseline, 89 participants were enrolled in the study. Four participants were excluded due to incomplete questionnaire responses, resulting in 85 participants completing the baseline evaluation. Among these 85 participants, all responded to the Epworth Sleepiness Scale (ESS), Pittsburgh Sleep Quality Index (PSQI), and Short Form-8 (SF-8) questionnaires. The Work Limitations Questionnaire (WLQ) was administered to 62 participants who were employed at the time of baseline visits.

Sixty-four participants initiated treatment for sleep disorders at our center and were eligible for follow-up evaluation. Among them, 38 participants completed the follow-up evaluation. However, some participants did not fully complete the WLQ at both time points, depending on their employment status: 8 participants did not respond to the WLQ at either baseline or follow-up, 2 participants responded to the WLQ at baseline but not at follow-up, and 5 participants responded to the WLQ only at follow-up. Thus, a total of 23 participants had complete WLQ data at both baseline and follow-up and were included in the longitudinal analysis (Figure 1). Longitudinal changes in daytime sleepiness measured by the ESS score, sleep quality measured by the PSQI, and health-related quality of life measured by SF-8 were analyzed as secondary outcomes.

Due to clinical scheduling constraints, patients typically waited 2-3 months before undergoing polysomnography, followed by an additional 2-3 weeks to receive their results. After initiating treatment, patients underwent a minimum of 3 months of treatment before the follow-up evaluation. As a result, the median duration from baseline to follow-up was 153 days (interquartile range: 140-199 days).

### Questionnaires

This study utilized validated Japanese versions of the following questionnaires: the ESS [8,9], PSQI [10,11], SF-8 [12,13], and WLQ [14–16]. The ESS is a questionnaire designed to measure sleep propensity in eight different situations [8,9]. In each situation, responders were asked to rate on a scale of 0-3, 0 as ‘would never doze’ and 3 as ‘high chance of dozing’. These scores are summed to obtain a total score ranging from 0 to 24, with higher scores indicating stronger subjective sleepiness. In general, ESS scores ≥ 11 are considered to indicate excessive daytime sleepiness (EDS) [17]. PSQI questionnaire measures sleep quality and disturbances; a global PSQI score of more than five is considered to have high sensitivity and specificity of distinguishing ‘poor’ sleepers [10,11]. The SF-8 is an abbreviated version of SF-36 (the 36-item health survey) and consists of eight questions that evaluate health-related quality of life (HRQOL)[12,13]. In the present study, physical and mental component scores (PCS and MCS) of the SF-8 were calculated according to the manual for users of the Japanese version of SF-8. Higher PCS and MCS scores indicate better HRQOL from the physical and mental health perspectives, respectively [13]. The WLQ is a questionnaire developed by Lerner that contains 25 questions originally designed to measure limitations due to health problems of employed individuals and health-related productivity losses, with its validity supported by a study comparing it to objective measures of work productivity. [14–16]. It consists of four subscales: a time management scale to measure handling time and scheduling demands; a physical demands scale to measure a person’s ability to perform physical demands; a mental-interpersonal demands scale to measure cognitive problems related to work and on-the-job social interactions; and output demands scale measuring diminished work quantity and quality. A higher WLQ score indicated higher presenteeism and productivity loss caused by health problems at work [18]. WLQ has been used to assess presenteeism in patients with various disorders, including low back pain, ankylosing spondylitis, and dry eye [19–21].

### Evaluation and treatment of sleep disorders

Evaluation of the sleep disorders was done mostly (96% of all participants) by sleep specialists certified by the Japanese Society of Sleep Research (S.T. and O.Y.) and the remaining others (4%) by a cardiologist (T.U.) supervised by sleep specialists. Diagnoses were made according to the International Classification of Sleep Disorders, third edition (ICSD-3), based on clinical history, polysomnography (PSG), multiple sleep latency test (MSLT), and sleep logs. Specific diagnostic criteria, such as an apnea-hypopnea index (AHI) threshold for OSA or sleep latency cutoffs for hypersomnia, followed standard guideline recommendations. While a full listing of diagnostic criteria is beyond the scope of this manuscript, we confirm that diagnoses adhered to established international standards.

Treatment for sleep disorders was initiated after a diagnosis was made, following practice guidelines from the American Academy of Sleep Medicine. The specific treatment approach varied by sleep disorder, as follows:

- Obstructive Sleep Apnea (OSA): The majority of patients were treated with continuous positive airway pressure (CPAP), while a small subset received oral appliances.
- Central disorders of hypersomnolence (e.g., narcolepsy, idiopathic hypersomnia): Treatment primarily involved modafinil or other wake-promoting agents, in addition to lifestyle guidance, including ensuring sufficient sleep duration and structured nap strategies.
- Insomnia: Patients were managed with hypnotic medications and lifestyle interventions based on cognitive behavioral therapy for insomnia (CBT-I) principles.
- Restless Legs Syndrome (RLS): Patients with iron deficiency received iron supplementation, while pharmacological treatment included dopaminergic agents or α2δ ligands.
- Circadian Rhythm Sleep-Wake Disorders (CRSWD): Management was based on sleep-wake logs, with interventions including lifestyle modifications, melatonin receptor agonists, and timed light exposure.
- Parasomnia: Treatment was individualized and included pharmacotherapy where appropriate, as well as management of coexisting sleep disorders that might contribute to parasomnic events. While these treatments followed guideline-based recommendations, minor adjustments were occasionally made according to patient preferences and the Japanese medical insurance system.

The time from diagnosis to follow-up evaluation was monitored, and follow-up questionnaires to assess treatment efficacy were conducted during clinic visits that took place at least three months after treatment initiation.

### Statistical analysis

Baseline data between the groups were compared using the Mann-Whitney U and chi-square tests.

In the longitudinal analysis, to ensure consistency in interpretation, all change scores were defined so that an improvement resulted in a positive value. For most variables, where a decrease indicated improvement, the change score was calculated as (pre-treatment value) − (post-treatment value). However, for the SF-8 physical component score and mental component score, where an increase indicated improvement, the change score was calculated as (post-treatment value) − (pre-treatment value). For most variables, which followed a normal distribution, statistical comparisons were conducted using paired t-tests, and 95% confidence intervals (CIs) for the mean were derived from the test results. Effect sizes were calculated using Cohen’s d. For variables where normality was not confirmed, the Wilcoxon signed-rank test was performed instead. Additionally, for these variables, the 95% CI for the median change was estimated using the bootstrap method (10,000 replications with replacement). The percentile method was applied, with the 2.5th and 97.5th percentiles of the bootstrapped distribution serving as the lower and upper bounds. The effect size r was calculated as r = Z / √n, where Z was derived from the Wilcoxon signed-rank test statistic.

To assess potential attrition bias, baseline characteristics were compared between participants who completed follow-up and those who were lost to follow-up. Differences between groups were evaluated using t-tests or Wilcoxon rank-sum tests for continuous variables, and chi-square tests for categorical variables.

We also conducted sensitivity analyses using two approaches: (1) a best-case / worst-case scenario and (2) the last observation carried forward (LOCF) method. In the best-case scenario, missing follow-up values were replaced with the 95th percentile of the observed distribution, assuming maximal treatment benefit. Conversely, in the worst-case scenario, missing follow-up values were replaced with the 5th percentile, assuming minimal or no treatment benefit. In LOCF analysis, missing follow-up values were imputed using their respective baseline values, assuming no further change. This approach provides a conservative estimate by minimizing the potential overestimation of treatment effects. For each sensitivity analysis, the same statistical tests used in the primary analysis were applied. Paired t-tests were conducted for normally distributed data, while the Wilcoxon signed-rank test was used for non-normally distributed data. Effect sizes were calculated using Cohen’s d for parametric tests and r for nonparametric tests.

All statistical analyses were performed using the SAS OnDemand for Academics. The version of Base SAS Software used in SAS OnDemand for Academics for this study is Custom version information: 9.4_M7. All P-values were two-tailed, and values of < 0.05 were regarded as statistically significant.

## RESULTS

### Baseline analysis

A total of 85 participants (mean age 40.2 years, 42% female) completed the baseline evaluation (Table 1).

**Table 1.** Characteristics of participants according to their diagnoses.

|  | Insomnia | SRBD | CH | CRSWD | Parasomnia | SRMD | No sleep disorder | Not definite | All |
| --- | --- | --- | --- | --- | --- | --- | --- | --- | --- |
| Number diagnosed | 7 | 42 | 27 | 1 | 1 | 3 | 3 | 1 | 85 |
| Age (years) | 40 (15.5) | 47.3 (12.2) | 29.7 (11.2) | In their 20s* | In their 60s* | 48.0 (18.5) | 30.7 (10.1) | In their 30s* | 40.2 (14.7) |
| Male | 2 (28.6) | 32 (76.2) | 11 (40.7) | 0 (0) | 0 (0) | 2 (66.7) | 2 (66.7) | 0 (0) | 49 (57.6) |
| ESS score | 12.9 (4.1) | 11.2 (5.8) | 16.0 (4.1) | 18 (-) | 2.0 (-) | 12.3 (2.5) | 10.7 (7.1) | 18 (-) | 12.9 (5.51) |
| ESS≥11 | 4 (57.1) | 21 (50.0) | 24 (88.9) | 1 (100) | 0 (0) | 2 (66.7) | 2 (66.7) | 1 (100) | 55 (64.7) |
| PSQI score | 14.7 (4.11) | 7.2 (2.92) | 7.4 (2.59) | 11.0 (-) | 4.0 (-) | 9.33 (2.89) | 6 (2.00) | 9 (-) | 7.98 (3.52) |
| SF-8 Physical component score | 42.7 (9.87) | 45.8 (9.66) | 49.4 (5.85) | 49.9 (-) | 53.1 (-) | 45.3 (3.19) | 55.1 (4.42) | 46.3 (-) | 47.1 (8.42) |
| SF-8 Mental component score | 38.9 (10.1) | 46.4 (7.29) | 45.0 (9.17) | 46.0 (-) | 51.0 (-) | 41.5 (7.49) | 39.5 (10.5) | 46.6 (-) | 45.0 (8.32) |
| Number with WLQ data | 4 | 33 | 21 | 0 | 0 | 2 | 2 | 0 | 62 |
| WLQ Time management | 52.5 (27.2) | 16.7 (20.0) | 26.8 (27.3) | NA | NA | 5 (0.0) | 15 (21.21) | NA | 22.0 (24.3) |
| WLQ Physical demands | 26.3 (25.0) | 24.4 (24.4) | 17.9 (18.7) | NA | NA | 12.5 (17.7) | 25 (11.8) | NA | 22.0 (21.9) |
| WLQ Mental-interpersonal demands | 44.5 (24.2) | 22.5 (18.6) | 30.7 (23.1) | NA | NA | 18.1 (5.89) | 11.1 (7.86) | NA | 26.2 (20.8) |
| WLQ Output demands | 38.8 (39.02) | 20.0 (23.8) | 30.4 (27.7) | NA | NA | 15 (14.1) | 2.5 (3.54) | NA | 24.0 (26.1) |

\*To prevent potential identification of individual participants, precise ages have been replaced with approximate age ranges.
Data presented as mean (standard deviation) or n(%).
SRBD: sleep-related breathing disorders, CH: central hypersomnia, CRSWD: circadian rhythm sleep-wake disorder, SRMD: sleep-related, movement disorders, ESS: Epworth Sleepiness Scale, PSQI: Pittsburgh Sleep Quality Index, WLQ: Work Limitations Questionnaire, NA: not available

All participants responded to ESS, PSQI, and SF-8. The WLQ was administered to 62 participants who were working at the time of baseline visits.

Table 1 presents the characteristics of the participants, based on their diagnoses. Diagnoses were classified according to ICSD-3, which means “central hypersomnia” (CH) category contains participants diagnosed not only with narcolepsy or idiopathic hypersomnia but also with insufficiency syndrome or long sleeper. The ‘Not definite’ category contained participants with inadequate data to make a diagnosis (e.g. canceling polysomnography). Ten participants were diagnosed with multiple sleep disorders, and in that case, the most clinically relevant diagnosis was chosen by the author (S.T.). The most common diagnosis was sleep-related breathing disorder (42 participants, 49%), and central hypersomnia was the second most common (27 participants, 32%). The average ESS score of the participants exceeded 11 in all diagnostic categories other than that with parasomnia and those diagnosed with no sleep disorder.

Table 2 shows a comparison of participants with or without EDS (defined as ESS ≥ 11 or not). Participants with EDS were significantly younger. Their MCS of SF-8 and sleep quality measured by PSQI were more impaired; and scores on presenteeism subscales from the WLQ regarding time management, mental-interpersonal demands, and output demands were significantly higher.

**Table 2.**
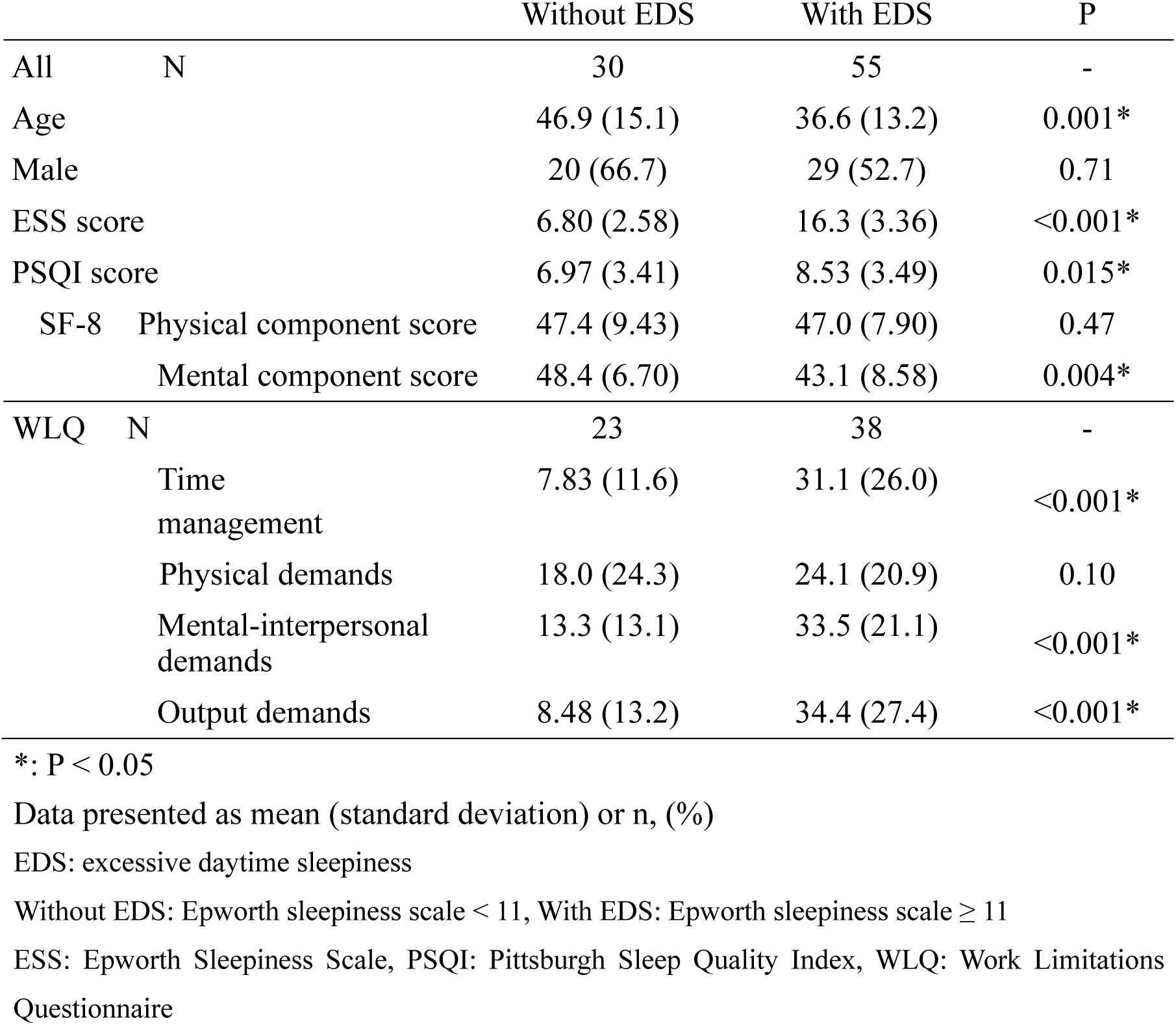
Comparison of participants with or without excessive daytime sleepiness in the baseline analysis.

### Longitudinal analysis

A total of 85 participants completed the baseline evaluation and 64 initiated treatment for sleep disorders at our center.

Among them, 38 participants completed the follow-up evaluation, with a mean duration from baseline to follow-up of 204 days (standard deviation: 119.6 days). Of these, 23 participants had complete WLQ data at both baseline and follow-up and were included in the final longitudinal analysis (Figure 1). Notably, 9 out of 10 participants who were diagnosed with multiple sleep disorders at baseline completed follow-up.

Table 3 shows a comparison of baseline characteristics between patients with and without follow-up data. Patients who did not complete follow-up evaluation were significantly younger (P =0.02), had a better SF-8 mental component score (P = 0.02), and exhibited better WLQ output demands scores (P = 0.02).

**Table 3.** Comparison of participants with or without follow-up data in the baseline analysis.

|  |  | With follow-up<br>data | Without follow-up<br>data | P |
| --- | --- | --- | --- | --- |
| All | N | 38 | 47 |  |
| Age |  | 43.8 (14.9) | 37.3 (13.9) | 0.02* |
| Male |  | 21 (55.3) | 28 (59.6) | 0.21 |
| ESS score |  | 13.6 (5.65) | 12.4 (5.39) | 0.33 |
| PSQI score |  | 8.47 (3.78) | 7.57 (3.28) | 0.37 |
| SF-8 | Physical component score | 46.7 (7.84) | 47.4 (8.93) | 0.33 |
|  | Mental component score | 42.9 (7.94) | 46.7 (8.33) | 0.02* |
| WLQ | N | 25 | 36 |  |
|  | Time management | 22.9 (22.5) | 21.9 (25.9) | 0.5 |
|  | Physical demands | 27.1 (24.3) | 18.2 (21.8) | 0.08 |
|  | Mental-interpersonal demands | 29.0 (18.3) | 23.7 (22.4) | 0.13 |
|  | Output demands | 30.8 (22.8) | 20.4 (27.8) | 0.02* |

\*: $P < 0.05$
Data presented as mean (standard deviation) or n, (%)
ESS: Epworth Sleepiness Scale, PSQI: Pittsburgh Sleep Quality Index, WLQ: Work Limitations Questionnaire

Table 4 shows the change in symptoms between the baseline and follow-up evaluation regarding ESS score, PSQI score, and SF-8 summary.

**Table 4.** Change in symptoms between the baseline and the follow-up evaluation regarding ESS score, PSQI score, and SF-8 summary.

|  | N | Baseline<br>(std) | Follow up<br>(std) | Change<br>of the<br>variables<sup>1</sup><br>(95%<br>CI)<sup>2</sup> | Effect<br>size<sup>3</sup> | P |
| --- | --- | --- | --- | --- | --- | --- |
| ESS score<sup>4</sup> | 38 | 13.6 (5.65) | 11.8 (5.43) | 1.79 (0-2) | 0.17 | 0.01* |
| PSQI score<sup>4</sup> | 38 | 8.47 (3.78) | 6.66 (2.74) | 1.82 (0-2) | 0.27 | <0.001* |
| SF-8 Physical component score | 38 | 46.7 (7.84) | 48.2 (7.73) | 1.00 (-1.12, 3.12) | 0.20 | 0.34 |
| SF-8 Mental component score | 38 | 42.9 (7.94) | 45.5 (8.92) | 2.54 (-0.83, 5.93) | 0.25 | 0.14 |

\*P <0.05
1. Change scores were defined so that improvement resulted in a positive value. Specifically, for variables where a decrease indicated improvement, the change score was calculated as (pre-treatment value) – (post-treatment value), while for variables where an increase indicated improvement, the change score was calculated as (post-treatment value) – (pre-treatment value).
2. The 95% confidence intervals (CIs) were calculated using different methods depending on normality. For data assuming normality (the majority of the dataset), CIs for the mean were derived from the paired t-test. For data where normality was not confirmed (as indicated in Footnote 3), CIs for the median were estimated using the bootstrap method.
3. For data assuming normality (the majority of the dataset), Cohen's d was used as the effect size. For data where normality was not confirmed (as indicated in Footnote 3), the effect size r was derived from the Wilcoxon signed-rank test.
4. Data where normality was not confirmed.
ESS: Epworth Sleepiness Scale, PSQI: Pittsburgh Sleep Quality Index, WLQ: Work Limitations Questionnaire

The ESS score (average −1.79 point, P = 0.01) and the PSQI score improved significantly (average −1.82-point, P < 0.001) after treatment, although SF-8 scores did not. Table 5 shows the changes in symptoms of the 23 participants who responded to the WLQ at both baseline and follow-up evaluations.

**Table 5.** Change in symptoms between the baseline and the follow-up evaluation in participants responding to all questionnaires including WLQ on both occasions.

|  |  | Baseline<br>(Std) | Follow up<br>(Std) | Change of<br>the<br>variables<sup>1</sup><br>(95% CI)<sup>2</sup> | Effect size<sup>3</sup> | P |
| --- | --- | --- | --- | --- | --- | --- |
| All | N | 23 | 23 | - | - | - |
| ESS score |  | 13.7 (5.70) | 11.9 (5.14) | 1.78 (2.91,<br>5.33) | 0.37 | 0.03* |
| PSQI score<sup>4</sup> |  | 8.61 (3.42) | 7.13 (2.20) | 1.48 (0, 2) | 0.20 | <0.001* |
| SF-8 | Physical<br>component<br>score | 47.2 (7.70) | 48.2 (7.47) | 1.00 (-1.12,<br>3.12) | 0.20 | 0.34 |
|  | Mental<br>component<br>score | 43.0 (5.90) | 46.4 (7.79) | 3.34 (0.13,<br>6.55) | 0.45 | 0.04* |
| WLQ | Time<br>management | 21.4 (22.1) | 15.2 (16.1) | 6.20 (-<br>1.02,13.4) | 0.37 | 0.09 |
|  | Physical<br>demands | 27.0 (22.9) | 17.3 (21.7) | 9.67 (-0.21,<br>19.6) | 0.42 | 0.04* |
|  | Mental-interpersonal<br>demands | 27.0 (17.3) | 21.2 (13.1) | 5.73 (-0.90,<br>12.3) | 0.37 | 0.086 |
|  | Output<br>demands | 29.4 (22.9) | 15.9 (16.5) | 13.5 (5.86,<br>21.1) | 0.77 | 0.001* |
| Baseline ESS <11 | N | 8 | 8 | - | - | - |
| ESS score |  | 7.50 (2.39) | 7.25 (2.76) | 0.25 (-2.22,<br>2.73) | 0.08 | 0.82 |
| PSQI score<sup>4</sup> |  | 7.25 (3.01) | 6.75 (1.91) | 0.5 (-0.5, 2) | 0.087 | 0.50 |
| SF-8 | Physical<br>component<br>score | 48.4 (4.37) | 50.9 (4.46) | 2.52 (-1.44,<br>6.47) | 0.53 | 0.18 |
|  | Mental<br>component<br>score | 47.1 (3.97) | 47.5 (6.89) | 0.36 (-3.40,<br>4.11) | 0.079 | 0.83 |
| WLQ | Time<br>management | 5.00 (7.07) | 9.38 (9.04) | -4.38 (-12.9,<br>4.11) | -0.43 | 0.26 |
|  | Physical<br>demands | 31.7 (30.0) | 21.7 (34.3) | 10.0 (-18.8,<br>38.8) | 0.29 | 0.44 |
|  | Mental-interpersonal<br>demands | 19.2 (12.3) | 16.6 (11.7) | 2.53 (-3.34,<br>8.40) | 0.36 | 0.34 |
|  | Output<br>demands | 16.9 (15.3) | 9.38 (8.21) | 7.50 (-2.74,<br>17.7) | 0.61 | 0.13 |
| Baseline ESS $\geq 11$ | N | 15 | 15 | - | - | - |
| ESS score |  | 17.00<br>(3.82) | 14.40 (4.32) | 2.60 (2.91,<br>6.27) | 0.65 | 0.02* |
| PSQI score<sup>4</sup> |  | 9.33 (3.50) | 7.33 (2.38) | 2.0 (0.5, 3) | 0.24 | 0.001* |
| SF-8 | Physical component score | 46.5 (9.08) | 46.7 (8.44) | 0.19 (-2.55,<br>2.93) | 0.039 | 0.88 |
|  | Mental component score | 40.9 (5.67) | 45.8 (8.40) | 4.93 (0.33,<br>9.52) | 0.59 | 0.04* |
| WLQ | Time management<sup>4</sup> | 30.2 (22.6) | 18.3 (18.4) | 11.8 (5, 20) | 0.28 | 0.01* |
|  | Physical demands | 24.4 (18.8) | 14.9 (11.6) | 9.50 (1.18-17.8) | 0.63 | 0.03* |
|  | Mental-interpersonal demands | 31.1 (18.4) | 23.7 (13.5) | 7.43 (-2.70,<br>17.6) | 0.41 | 0.14 |
|  | Output demands | 36.0 (23.9) | 19.3 (18.9) | 16.7 (14.3,<br>30.8) | 0.85 | 0.005* |

\*P < 0.05
1. Change scores were defined so that improvement resulted in a positive value. Specifically, for variables where a decrease indicated improvement, the change score was calculated as (pre-treatment value) – (post-treatment value), while for variables where an increase indicated improvement, the change score was calculated as (post-treatment value) – (pre-treatment value). 2. The 95% confidence intervals (CIs) were calculated using different methods depending on normality. For data assuming normality (the majority of the dataset), CIs for the mean were derived from the paired t-test. For data where normality was not confirmed (as indicated in Footnote 3), CIs for the median were estimated using the bootstrap method. 3. For data assuming normality (the majority of the dataset), Cohen's d was used as the effect size. For data where normality was not confirmed (as indicated in Footnote 4), the effect size *r* was derived from the Wilcoxon signed-rank test. 4. Data where normality was not confirmed.
ESS: Epworth Sleepiness Scale, PSQI: Pittsburgh Sleep Quality Index, WLQ: Work Limitations Questionnaire
Without EDS: Epworth sleepiness scale < 11, With EDS: Epworth sleepiness scale $\geq 11$

All subscales of the WLQ improved significantly compared to baseline (time management: P = 0.049, physical demands: P = 0.035, mental-interpersonal demands: P =0.035, output demands: P < 0.001; Figure 2); as did the ESS score, PSQI score, and mental component summary of the SF-8. When we divided participants based on baseline ESS (ESS ≥ 11, ESS <11) and repeated the analysis, mental component scores of SF-8 and three subscales of WLQ (time management, physical demands, and output demands) significantly improved in patients with high baseline ESS, while no measures of PSQI, SF-8, and WLQ significantly changed in participants with low baseline ESS.

**Figure 2:**
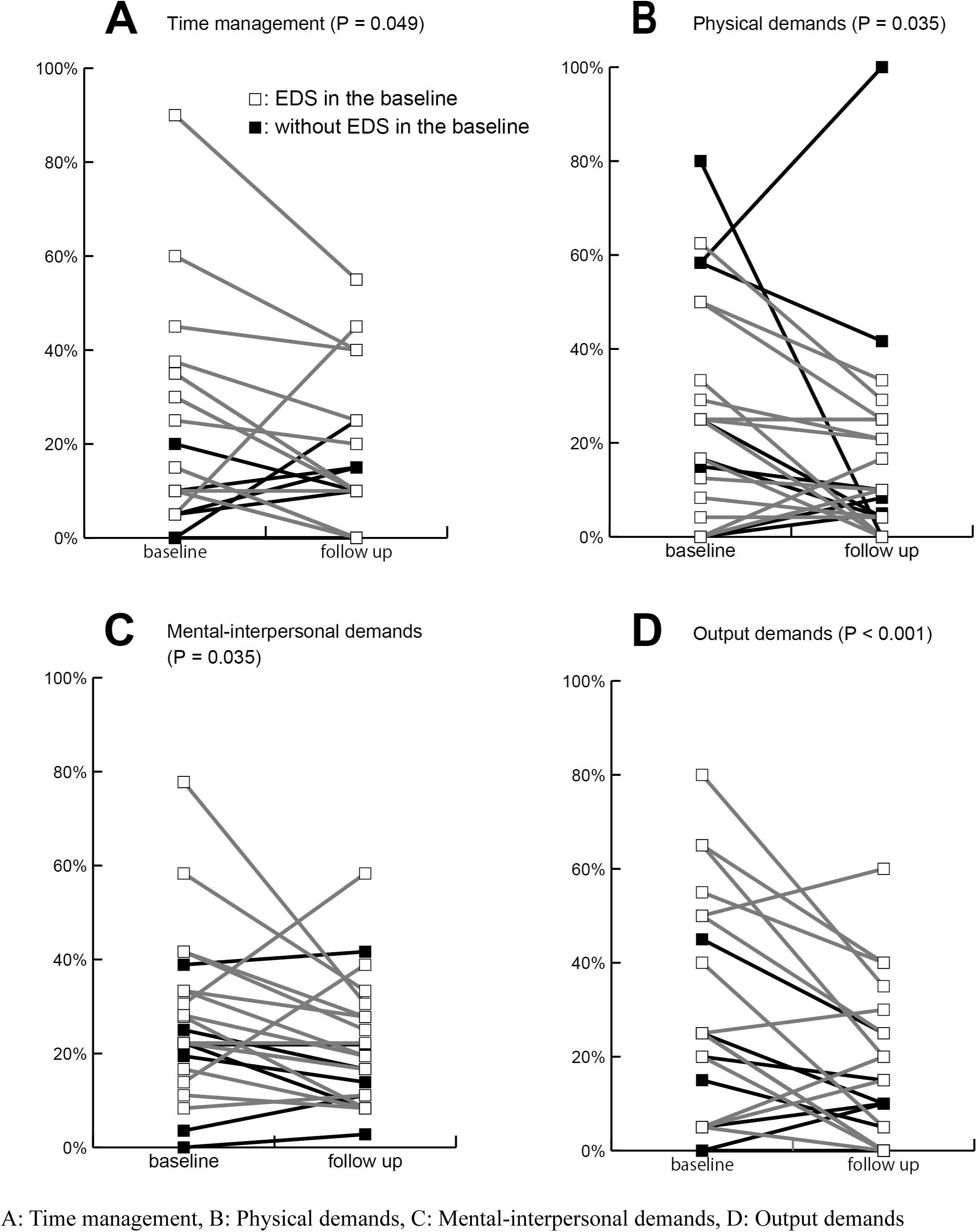
Changes observed in each participant regarding subscales of Work Limitations Questionnaire between the baseline and the follow-up.

To assess the robustness of the findings, sensitivity analyses were conducted using the best-case / worst-case scenario (Table 6). Best-case scenario analysis showed statistically significant improvements in all variables after treatment. In contrast, the worst-case scenario resulted in a more extreme outcome, where no variables that were originally significant retained their significance. Furthermore, in many cases, variables exhibited a significant worsening in the worst-case assumption. However, ESS scores and PSQI scores in the overall analysis, and PSQI scores, the mental component score of SF-8, and three WLQ subscales (time management, physical demands, and output demands) in the group with EDS did not show significant worsening even under the worst-case assumption.

**Table 6:**
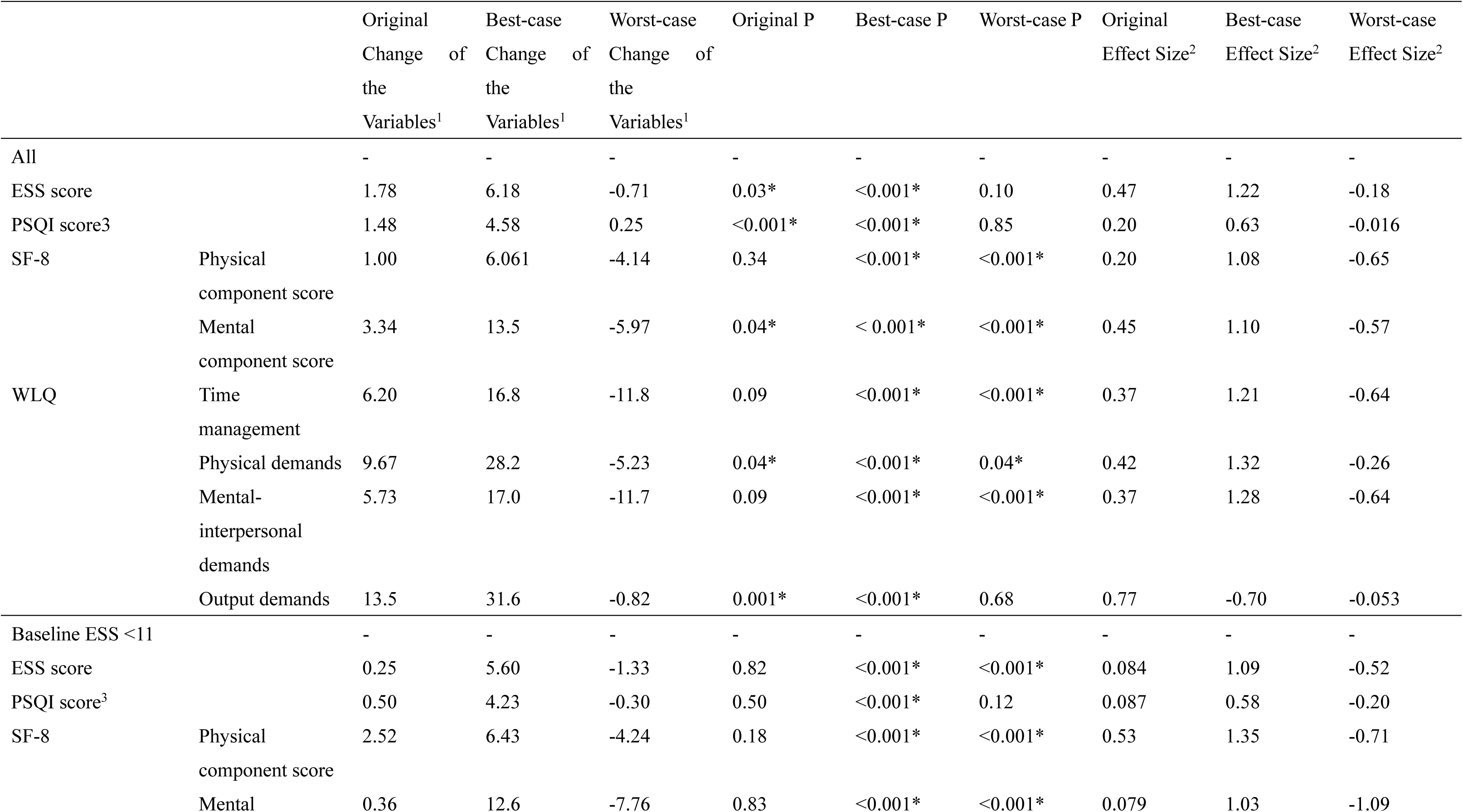
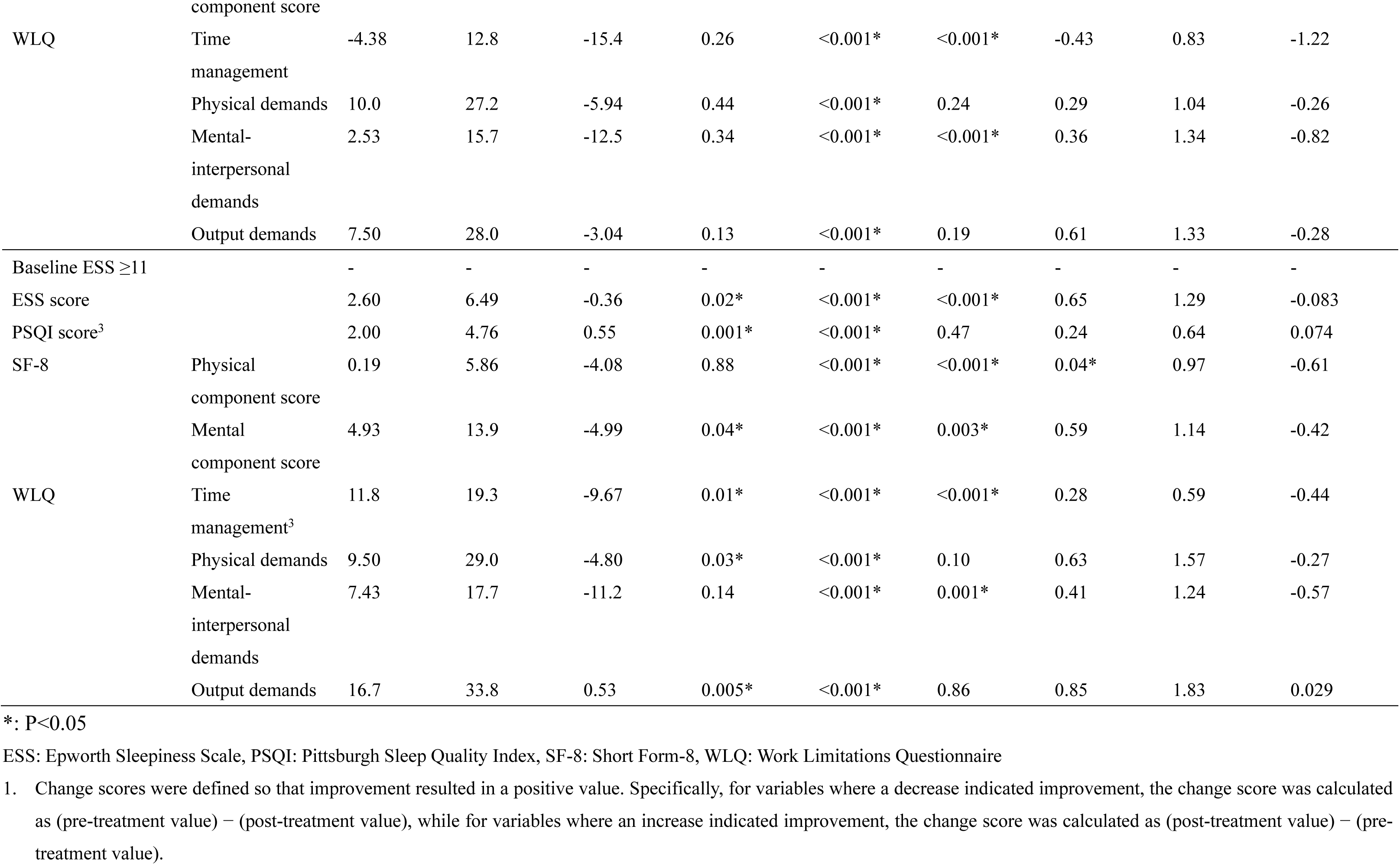
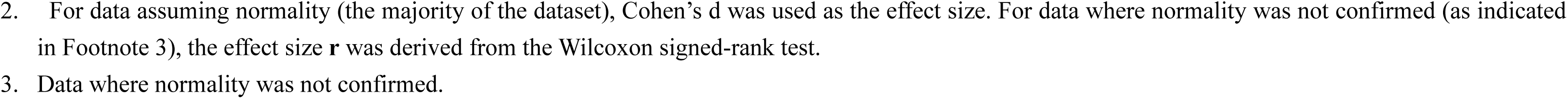
Sensitivity Analysis: Best-Case and Worst-Case Scenarios.

Since the best/worst-case scenarios tended to produce overly extreme results, we performed an additional sensitivity analysis using the LOCF method, which assumes no change for missing values (Table 7). While effect sizes were lower in the LOCF analysis compared to the primary analysis, the overall results remained consistent. In the full dataset, variables ESS scores, PSQI scores, and two WLQ subscales (physical demands and output demands) showed significant improvement after treatment, even under the LOCF assumption. Among participants with EDS, ESS scores, PSQI scores, and three subscales of WLQ (time management, physical demands, output demands) retained statistical significance.

**TABLE 7:**
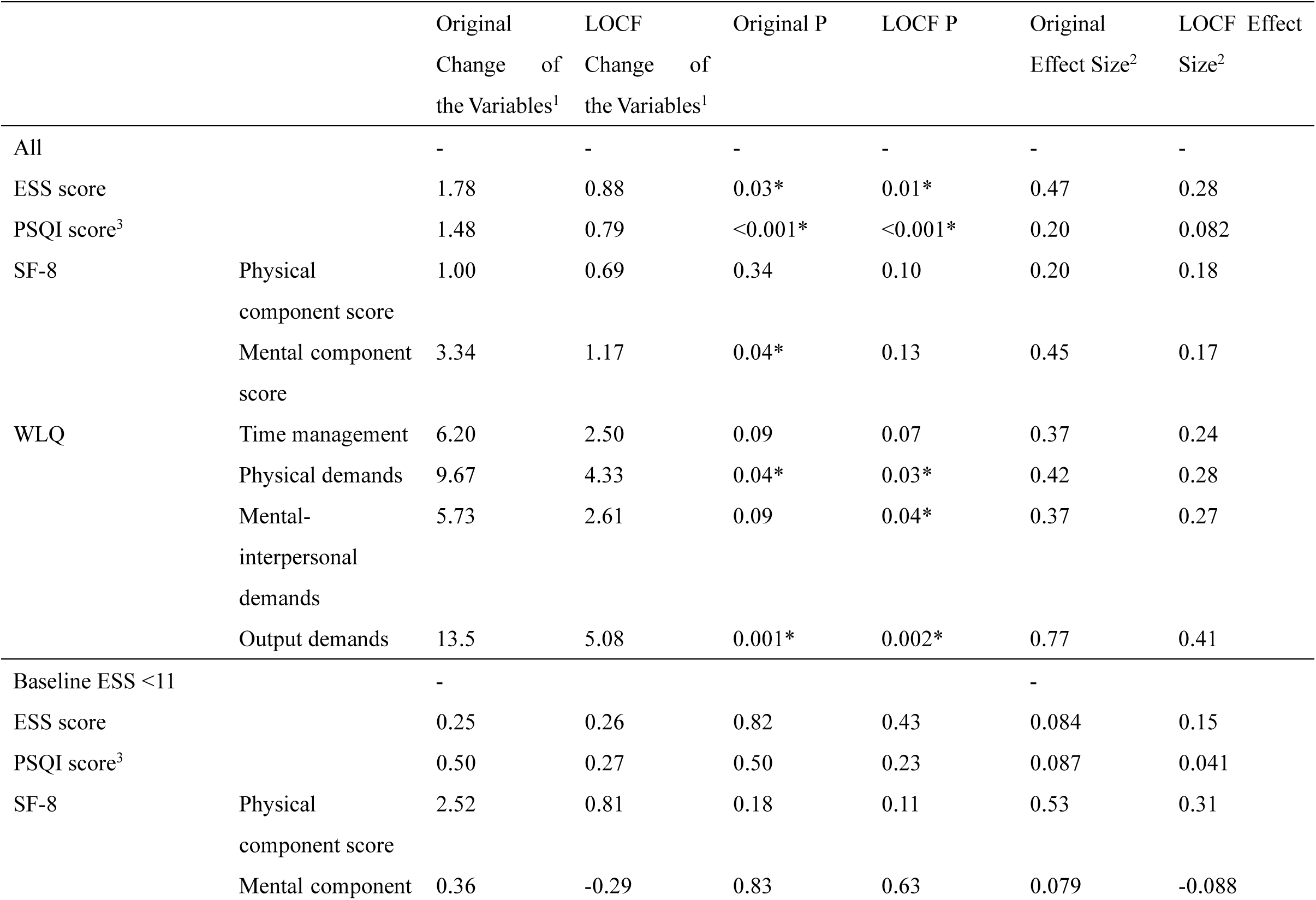
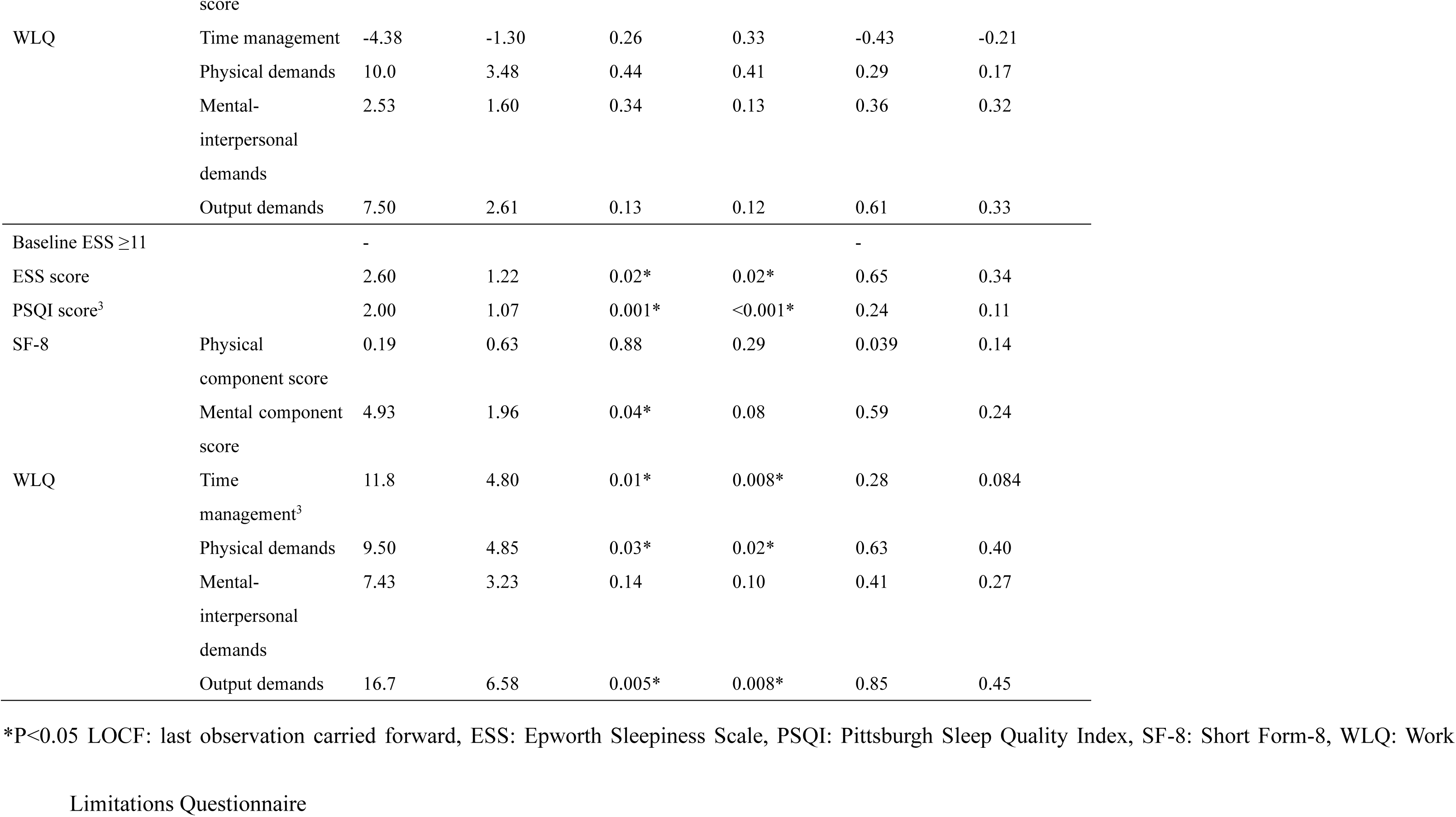
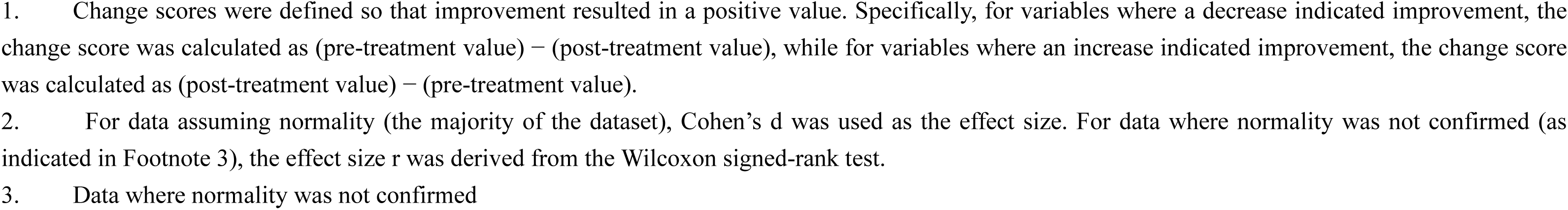
Sensitivity Analysis Using Last Observation Carried Forward (LOCF)

## DISCUSSION

### Summary of findings

In the baseline analysis, participants with excessive daytime sleepiness (EDS) exhibited greater impairments in three WLQ subscales (time management, mental-interpersonal demands, and output demands) compared to those without EDS. In the longitudinal analysis, all WLQ subscales significantly improved after three months of sleep disorder treatment, a trend that remained robust even when the analysis was limited to participants with EDS. These findings suggest that while work productivity is more impaired in sleep disorder patients with EDS, their impairments were observed to improve following treatment.

### Comparison with previous studies

Various sleep disorders involving EDS have been associated with productivity loss. Dean et al. reported that excessive sleepiness (ESS ≥10) correlated with impaired work productivity and lower health-related quality of life (HRQOL) in a large U.S.-based survey [6]. Stepnowsky et al. found that OSA with EDS was linked to reduced productivity [5], and similar findings have been reported in patients with narcolepsy [7]. Our baseline findings align with these previous studies.

Presenteeism, defined as reduced workplace productivity due to health conditions, is expected to improve with effective treatment. Prior studies have shown that presenteeism improves with treatment in patients with diabetes [22], rheumatoid arthritis [23], and major depressive disorder [24]. In sleep medicine, cognitive behavioral therapy for insomnia [25] and solriamfetol for OSA-related sleepiness [26] have been reported to enhance productivity. Shimura et al. showed that individuals with eveningness preference tended to have higher levels of presenteeism, though later wakeup time with those with eveningness tendency was associated with lower presenteeism, suggesting that appropriate wakeup times for eveningness people may help reduce presenteeism [27]. Our study observed presenteeism improvements across different sleep disorders following guideline-based treatments (e.g., CPAP for SAS, modafinil for hypersomnia). While many studies focus on specific sleep disorders, our study included patients with diverse sleep disorders to better reflect real-world clinical practice, where patients seek treatment based on symptoms rather than predefined diagnostic categories. For example, 10 participants (11.8%) in our study had multiple sleep disorders, making strict categorization challenging. Instead of stratifying by diagnosis, we aimed to examine whether individuals seeking treatment for sleep-related concerns experienced improvements in work productivity following treatment, supporting the generalizability of these findings to routine clinical practice.

Excessive daytime sleepiness due to sleep disorders can have severe employment consequences. Patients with narcolepsy in Denmark exhibit higher unemployment rates than controls [28], and Japanese studies indicate that individuals with central hypersomnia experience job dismissals and turnover [29]. Such employment difficulties extend beyond hypersomnia, affecting patients with OSA as well [30]. If reduced work productivity due to sleep disorders contributes to these challenges, our findings suggest that encouraging timely evaluation and treatment could potentially mitigate productivity-related employment risks.

### Implications for workplace interventions

While significant improvements in work productivity were observed, WLQ scores remained above zero, indicating that presenteeism did not fully resolve. This suggests that while treatment leads to notable improvements, residual deficits persist. A comprehensive approach integrating clinical treatment with workplace strategies could further support individuals managing sleep disorders. While our study did not directly evaluate workplace interventions, previous research suggests that strategies such as cognitive behavioral therapy for insomnia [25] programs and flexible work schedules [27] may help mitigate sleep-related impairments. Future studies should investigate the potential role of workplace adaptations in supporting employees with sleep-related impairments. Strengths and limitations

This study has several strengths. First, the longitudinal design of this study enabled the assessment of within-subject changes in work productivity and sleep-related symptoms over time, although the high attrition rate necessitates a cautious interpretation of these findings. Second, we included patients with various sleep disorders rather than focusing on a single condition. Since many patients have overlapping sleep disorders, a diagnosis-based approach may oversimplify real-world clinical scenarios. By including a diverse patient population, our study aimed to capture the overall impact of sleep disorder treatment on work productivity, independent of specific diagnostic labels. Third, the study was conducted at a tertiary sleep center, where patients underwent comprehensive diagnostic evaluations by sleep specialists, ensuring accurate sleep disorder diagnoses and the implementation of guideline-based treatments. Fourth, validated and widely recognized assessment tools-including the Epworth Sleepiness Scale (ESS), Pittsburgh Sleep Quality Index (PSQI), Short Form-8 (SF-8), and Work Limitations Questionnaire (WLQ)-were used, enhancing comparability with previous studies. Finally, while many studies have examined the health consequences of sleep disorders, their impact on work productivity remains underexplored. By utilizing the WLQ, which has been validated against objective measures of work productivity [15], this study provides valuable insights into the occupational implications of sleep disorder treatment.

This study has several limitations. First, the high attrition rate (55%) introduces potential bias, as participants who completed follow-up tended to have worse baseline health indicators than those lost to follow-up. While sensitivity analyses were conducted to assess the impact of missing data, the results should be interpreted with caution, particularly given that the worst-case scenario analysis did not yield statistically significant results.

Second, selection bias may have influenced the findings, as participants were recruited from a tertiary sleep center and may not represent the broader population of individuals with sleep disorders. Additionally, patients with more severe or persistent symptoms were more likely to remain in follow-up, while those with milder conditions were more likely to discontinue, potentially leading to an overrepresentation of individuals with greater impairment. Third, all outcome measures were based on self-reported questionnaires, which are subject to recall and reporting bias. Future research should incorporate objective measures, such as actigraphy or employer-reported work performance data, to validate these findings.

Fourth, variability in follow-up timing (median: 153 days, IQR: 140–199 days) introduces potential measurement inconsistencies. Although this reflects real-world clinical scheduling and patient adherence patterns, standardized follow-up intervals in future studies would help mitigate this issue.

Fifth, participants received heterogeneous treatments based on their specific diagnoses and clinical needs. While this reflects standard clinical practice, condition-specific analyses were not feasible due to the limited sample size. Future research with larger cohorts should explore diagnosis-specific treatment effects.

## CONCLUSION

In conclusion, our findings suggest that work productivity in patients with sleep disorders may improve following a few months of treatment. This trend was observed across different sleep disorder diagnoses, although the extent of improvement varied among individuals. Seeking appropriate treatment for sleep disorders may improve presenteeism, but further research is needed to confirm these findings. The relationship between sleep disorder treatment and employment status remains an important area for future investigation, particularly in assessing whether improved sleep health translates into better job retention and occupational functioning.

## Acknowledgments

The corresponding author is currently affiliated with Department of Public Health, Juntendo University. This study was conducted at Ehime University.

The authors would like to thank Editage (www.editage.jp) for English language editing.

## Abbreviations

OSA: obstructive sleep apnea
ESS: Epworth Sleepiness Scale
PSQI: Pittsburgh Sleep Quality Index
WLQ: Work Limitations Questionnaire
ESS: excessive daytime sleepiness
HRQOL: health-related quality of life
PCS: physical component scores
MCS: mental component scores
SF-8: Short Form-8 Health Survey

## Declarations

### Funding

The results reported herein correspond directly to the specific aims of KAKENHI Grant Number JP 17K15746 from the Japan Society for the Promotion of Science.

### Competing Interests

The authors have no competing interests to declare that are relevant to the content of this article.

### Ethics Approval

The questionnaire and methodology for this study was approved by the Institutional Review Board of the Ehime University Hospital (No.1709002).

All procedures complied with the Declaration of Helsinki and relevant national regulations/guidelines.

### Data Availability

The participants in this study provided consent to participate under the condition that the data would not be publicly available. Therefore, the data are not available for public access.

### Consent to Participate

Informed consent was obtained from all individual participants included in the study.

### Author Contributions

S.T. contributed to the conception of the study; acquisition, analysis, and interpretation of data; and writing of the initial version of the manuscript. T.U., A.K., H.S., Y.F., and T.K. contributed to data collection. Y.O. contributed to the conception of the study, as well as data acquisition, analysis, and interpretation.

## Notes

### Competing Interest Statement

The authors have declared no competing interest.

### Author Declarations

Ethics Approval: The questionnaire and methodology for this study was approved by the Institutional Review Board of the Ehime University Hospital (No.1709002).

